# Genome-wide investigation of DNA methylation in congenital adrenal hyperplasia

**DOI:** 10.1101/19008524

**Authors:** Leif Karlsson, Michela Barbaro, Ewoud Ewing, David Gomez-Cabrero, Svetlana Lajic

**Author notes:** **Corresponding author:** Correspondence should be addressed to Dr. Leif Karlsson, Department of Women’s and Children’s Health, Paediatric Endocrinology Unit (QB83), Karolinska University Hospital, SE-171 76 Stockholm, Sweden, or to. **Disclosure statement:** The authors have nothing to disclose.

## Abstract

**Background:** Patients with congenital adrenal hyperplasia (CAH) are at risk of long-term cognitive and metabolic sequelae with some of the effects being attributed to the chronic glucocorticoid treatment that they receive. This study investigates genome-wide DNA methylation in patients with CAH to determine whether there is evidence for epigenomic reprogramming as well as any relationship to patient outcome.

**Methods:** We analysed CD4+ T cell DNA from 28 patients with CAH (mean age=18.5 ±6.5 years [y]) and 37 population controls (mean age=17.0 ±6.1 y) with the Infinium-HumanMethylation450 BeadChip array to measure genome-wide locus-specific DNA methylation levels. Effects of CAH, phenotype and *CYP21A2* genotype on methylation were investigated as well as the association between differentially methylated CpGs, glucose homeostasis, blood lipid profile and cognitive functions. In addition, we report data on a small cohort of 11 patients (mean age=19.1, ±6.0 y) with CAH who were treated prenatally with dexamethasone (DEX) in addition to postnatal glucocorticoid treatment.

**Results:** We identified two CpGs to be associated with patient phenotype: cg18486102 (located in the *FAIM2* gene; rho=0.58, adjusted p=0.027) and cg02404636 (located in the *SFI1* gene; rho=0.58, adjusted p=0.038). cg02404636 was also associated with genotype (rho=0.59, adjusted p=0.024). Higher levels of serum C-peptide was also observed in patients with CAH (p=0.044). Additionally, levels of C-peptide and HbA1c were positively correlated with patient phenotype (p=0.044 and p=0.034) and genotype (p=0.044 and p=0.033), respectively. No significant association was found between *FAIM2* methylation and cognitive or metabolic outcome. However, *SFI1* TSS methylation was associated with fasting plasma HDL cholesterol levels (p=0.035).

**Conclusion:** In conclusion, higher methylation levels in CpG sites covering *FAIM2* and *SFI1* were associated with disease severity. Hypermethylation in these genes may have implications for long-term cognitive and metabolic outcome in patients with CAH.

## 1. Introduction

Congenital adrenal hyperplasia (CAH) due to 21-hydroxylase deficiency is one of the most common inborn errors of metabolism [1]. Ten common *CYP21A2* mutations are responsible for 95% of all CAH cases, where the phenotype is dictated by the less severe allele. The clinical manifestations of CAH include a broad spectrum of phenotypes and generally a good correlation exists between genotype and phenotype [2]. In the most severe salt-wasting form of CAH (SW CAH) the synthesis of both cortisol and aldosterone is severely affected. Indeed, a young infant will succumb to disease and then death in a salt-losing crisis if treatment is not initiated within the first weeks of life. In the simple-virilising form of CAH (SV CAH) aldosterone production is sufficient to avoid salt loss during basal conditions. Girls affected with classic forms of CAH are exposed to high levels of adrenal androgens prenatally and are born with virilised external genitalia. Dexamethasone (DEX) can be administered to pregnant mothers at risk of having a child with classic CAH to prevent the virilisation of girls with classic CAH already *in utero* [3].

The mild, non-classic form (NC CAH) is associated with signs of androgen excess from late childhood, but it may escape diagnosis until adulthood [4-6]. Postnatally, patients with CAH are treated with life-long glucocorticoid (GC) replacement therapy; for classic CAH, treatment with fludrocortisone is necessary additional to GCs [1]. During different periods throughout life, it may be difficult to achieve perfect dosing of GC replacement therapy, which ultimately leads to either over- or under-treatment [7].

With the implementation of neonatal screening programmes for CAH and continuous improvement in clinical care, CAH has become a lifelong chronic disease. Over time, patients with CAH are at risk of metabolic and cognitive sequelae later in life [8-11]. They typically present with obesity, insulin resistance and altered lipid profiles [7, 8, 12]. For cognitive outcome, while results are conflicting, the overall evidence suggests that patients with CAH are at risk of impaired executive functions and, more specifically, working memory [9-11, 13, 14]. In our current Swedish cohort, adult patients show deficits in executive functioning [9] as well as alterations in brain structures associated with the working memory network [15]. Our younger cohort of children with CAH performed equally to population controls regarding cognitive functions [16].

Moreover, mental health studies indicate that women and men with CAH have an increased risk of being diagnosed with a psychiatric disorder [17, 18].

Outcome may also depend on clinical phenotype and/or genotype, where patients with SW CAH have more difficulties than patients with milder phenotypes [8, 9, 19, 20]. While the exact causes of each outcome cannot always be identified, most commonly they are attributed to the GC replacement therapy and the inherent difficulties in mimicking the circadian cortisol rhythm. However, prenatal androgen exposure, prenatal GC deficit and postnatal adrenal crises may also affect long-term patient outcome.

In addition, prenatal treatment with DEX may have an impact on patient outcome. Previously, we have identified cognitive deficits in first-trimester DEX-treated children not having CAH. These effects seem to be sex-dimorphic because exposed girls were more affected than exposed boys [21]. In one of our recent reports we showed that women with CAH who were treated with DEX during the entire gestational period had broad deficits in cognition similar to what we observed in non-CAH girls during childhood [9].

Epigenetic mechanisms (such as DNA CpG methylation and histone modification) play a key role in the development of different diseases [22, 23]. The DNA methylation profile is set during development but does not remain stable throughout life. Changes in methylation have been associated with human disease, environmental changes, changes in physiological activity [24] and even changes in the social environment [25, 26]. Indeed, we have identified broad changes in DNA methylation measured in peripheral CD4+ T-cells from children not having CAH and exposed to prenatal DEX treatment during the first-trimester [27]. We may therefore postulate that DNA methylation is altered in patients with CAH and that it may be a potential mediating mechanism exerting influence on the long-term outcome.

This study investigates if epigenetic modification of DNA occurs in patients with CAH, and whether the methylation status is associated with the severity of the disease (clinical phenotype and genotype) in a unique and well-characterized group of patients. We hypothesize that DNA methylation is a stable fingerprint of the effects of CAH during embryonic development and later in life and that changes in CpG methylation are associated with metabolic and cognitive outcome. We also investigate whether prenatal DEX treatment generates long-lasting effects on CpG methylation profiles in a small but unique group of patients with CAH that have been exposed to such therapy either during the first trimester (CAH boys) or during the entire gestational period (CAH girls). To achieve this end we isolated CD4+ T cells from children and adults with CAH and from controls from the general population for comparison of DNA methylation levels and metabolic and cognitive outcome.

## 2. Methods

### 2.1 Participants

The study is part of a larger study investigating the effects of pre- and postnatal treatment in patients with CAH (for details, see Wallensteen *et al. [21]*). Here we investigate the effects of pre- and postnatal GC treatment on whole genome methylation in patients with CAH (vs. controls form the general population) and whether changes in CpG methylation are associated with disease severity, and cognitive and metabolic outcome. Written informed consent of all participants or their parents was obtained and the study was approved by the Regional Ethics Committee of Stockholm (dnr. 99-153).

In total, 76 participants were included in the study: 28 patients (12 females and 16 males) with CAH (mean age=18.5 years [y], ±6.5 y), 11 patients (7 females and 4 males) with CAH treated prenatally with DEX (mean age=19.1 y, ±=6.0 y) and 37 control subjects from the general population (18 females and 19 males, mean age=17.0 y, ±=6.1 y). Age did not differ significantly between groups (all ps>.05). Two patients with CAH had NC CAH, 13 had SV CAH and 24 had the SW phenotype. GC treatment in patients with CAH (not prenatally DEX-treated) were treated with 13.7 ±4.2 mg/m2 (HC equivalent dose) of GCs while prenatally treated patients received 13.0 ±3.0 mg/m2, the average dose did not differ significantly between groups (p>.05). The vast majority of the patients were diagnosed via the Swedish national neonatal screening programme for CAH.

### 2.2 Procedures

All participants were instructed not to eat after midnight the night before the visit. Blood samples for methylation analysis and analysis of glucose homeostasis and lipid profiles were collected in the morning. The participants’ height and weight were measured after sample collection. Blood (B) glucose, serum (S) insulin, S-C-peptide, B-HbA1c, plasma (P) triglycerides, P cholesterol, P HDL cholesterol and P LDL cholesterol were analysed at the accredited clinical chemistry laboratory at the Karolinska University Hospital. A detailed description of the procedures and tests used to assess cognitive performance is given in Karlsson *et al*, [9] and Wallensteen *et al. [21]*. Here, we included estimates of general intelligence (WISC-III/WAIS-IV, Matrices and Vocabulary) and executive functions (WISC-III/WAIS-IV, Coding, Digit Span and Stroop Interference) [28-30]. Before the analyses, raw scores from the neuropsychological assessments were transformed into scaled scores (mean=10, SD=3) based on age-specific Swedish norms for subtests from the Wechsler Scales. Raw scores from the Stroop Test were transformed into T scores (mean=50, SD=10) according to the North American norms [30].

### 2.3 Isolation of T-cells and flow cytometry

Peripheral blood mononuclear cells (PBMCs) were separated by density centrifugation from 50 ml of whole blood per participant. CD4+ T cells were isolated from the PBMCs using magnetic activated cell sorting (MACS, Miltenyi Biotech) and stored at - 80°C. The purity of CD4+ cell populations was evaluated by immune-phenotyping using two-colour antibody panels. Data were acquired and analysed using the Cyan ADP Analyzer (Summit 4.3, Beckman Coulter). T cell population purity was 94.9% (SD 3.1). For a more detailed description of T cell isolation and flow cytometry, see Reinius *et al*. [31]

### 2.4 DNA extraction, bisulphite treatment and DNA methylation measurements using the 450K Beadchip Array

Genomic DNA was isolated from T cell pellets using the QiAmp DNA Mini Kit (Qiagen) according to the manufacturer’s instructions. DNA concentration was measured using the Qubit 2.0 (Invitrogen). Bisulphite treatment was performed with the EZ-96 DNA Methylation Kit (Zymo Research) and DNA methylation measurements were done using the Illumina Infinium Human Methylation450 BeadChip Array (450k array, Illumina). Samples were analysed in two batches as part of a larger project investigating the epigenetic effects in individuals treated with DEX during fetal life and not having CAH [32] and in individuals with CAH who also receive treatment with glucocorticoids postnatally (the present study). Samples from individuals with CAH, prenatally treated participants and controls were distributed randomly on the arrays. Technical validation of the 450k arrays was performed using bisulphite pyrosequencing in the study investigating the effects of prenatal DEX given during the first trimester [32] and technical validation was thus not performed again in the present investigation.

### 2.5 DNA methylation quality control and data processing

The 450k array was used to determine DNA methylation levels at 480 000 CGs across the genome. The data analysis was performed in R and raw data were pre-processed using the lumi package [33, 34]. Three samples (all controls) were discarded during the quality control phase because of poor genome-wide correlation with other samples and an aberrant distribution of β values. In addition, the following probes were excluded during the pre-processing of the analysis: (i) probes located on the Y and X chromosomes, (ii) probes with a single nucleotide polymorphism (SNP) located within three base pairs of the interrogated CpG site in order to exclude false positive probes caused by genetic variations and (iii) CpG probes with poor detection p-values (*p*>0.01) [35].

After filtering, 395 462 probes remained. Estimations of β values for the probes were performed using a previously described three-step pipeline [36]. Batch effects were identified and their effect quantified using principal component analysis. Batch effects were subsequently corrected using the ComBat function from the sva Bioconductor package [37]. Finally, based on the assumption that most of the CAH-associated DNA methylation changes would be relatively small and that, while using all available samples, our sample size of patients and controls is limited, we choose to analyse highly variable probes; we selected probes whose interquartile range, after transforming β values into M values [34, 38], was >0.5. As a result, we selected 29 351 CpG sites for the association analysis.

### 2.6 DNA methylation differential analysis

Two analyses were conducted to study the association between DNA methylation and CAH. In the first analysis a linear model was generated for each CpG site for which the predictive variables for DNA methylation were: group (CAH-DEX or CAH), age, sex and group interaction with sex (CAH-DEX x sex) to investigate methylation between prenatally treated patients with CAH vs. those with CAH not prenatally treated. The analysis and linear models were generated separately for each sex due to the difference in DEX-treatment length. In the second analysis, a similar methodology was applied but included only patients with CAH not prenatally treated and population controls. To estimate the significance of each CpG site for each respective analysis we computed a permutation-based *p*-value in which 10 000 permutations were performed over the M-values for all CpG sites. A false discovery rate (FDR) was computed using a non-parametric method described elsewhere [39]. CpGs with a FDR <0.05 were considered significant.

### 2.7 DNA methylation quantitative trait analysis

We further sought to investigate the correlations between methylation and participant phenotype and *CYP21A2* genotype. Phenotype groups were defined and ranked by severity as: control, SV CAH and SW CAH to create three groups for the correlation analysis.

Genotypes were grouped based on the severity of the mildest mutated *CYP21A2* allele to create four groups for the correlation analysis. The groups were defined and ranked as wt (control), B (n=10, p.I172N), A (n=10, G291S, p.R356Q and I2 Splice) and null (n=7, no enzyme activity, including complete gene deletion, I7 Splice and p.R356W). The genetic status of the controls was not known but their mildest allele was assumed to be wt. Only patients with CAH not exposed to prenatal DEX were included in this analysis. One NC patient was excluded in that the NC phenotypic group included only this single patient.

Next, confounding effects of sex and age were regressed out from the methylation data using a linear model and the residual values obtained after correction were applied for correlation to either phenotype or genotype using Spearman’s non-parametric correlation. To estimate the significance of each CpG site for each respective analysis we computed a permutation-based *p*-value in which 10 000 permutations were performed over the residuals from the linear model corrected for sex and age. We selected significant CpG sites whose correlation between methylation levels and phenotype or genotype had a FDR of <0.05.

### 2.8 Analyses of patient outcomes

Group analysis of height, weight, BMI, glucose homeostasis, blood lipids and cognitive performance was performed using multiple linear regressions with CAH, age, sex and the CAH x sex interaction as predictors where appropriate (excluding age for cognition as the data were already age-corrected). We also performed non-parametric correlations to investigate the relationship between patient phenotype and *CYP21A2* genotype with metabolic and cognitive outcome. The correlation analyses were performed by correcting metabolic outcome data for age and sex and cognitive data for sex in a linear model. The residual values obtained after correction were applied for correlation to either phenotype or genotype using Spearman’s non-parametric correlation.

Associations between methylation and previously described clinical outcomes were performed using multiple linear regressions with β-values, age, sex and β-values x sex interaction as predictors (again excluding age for cognitive outcome data). Associations with a nominal *p*<0.05 were considered significant for all analyses. All statistical analyses were performed in R.

## 3. Results

### 3.1 Differential methylation analysis

No significant CpG sites were identified in the analysis investigating the effect of prenatal DEX treatment or in the analysis comparing CAH patients with population controls.

### 3.2 DNA methylation quantitative trait analyses

In the quantitative trait analyses phenotype correlated significantly with two CpG sites: cg18486102 (*rho*=0.58, adjusted *p*=0.027) and cg02404636 (*rho*=0.58, adjusted *- p*=0.038). cg02404636 also significantly correlated with genotype (*rho*=0.59, adjusted *- p*=0.024). Methylation (β-values) for both CpGs increased with the severity of the disease (genotype and phenotype) (Figure 1A and C).

**Figure 1.**
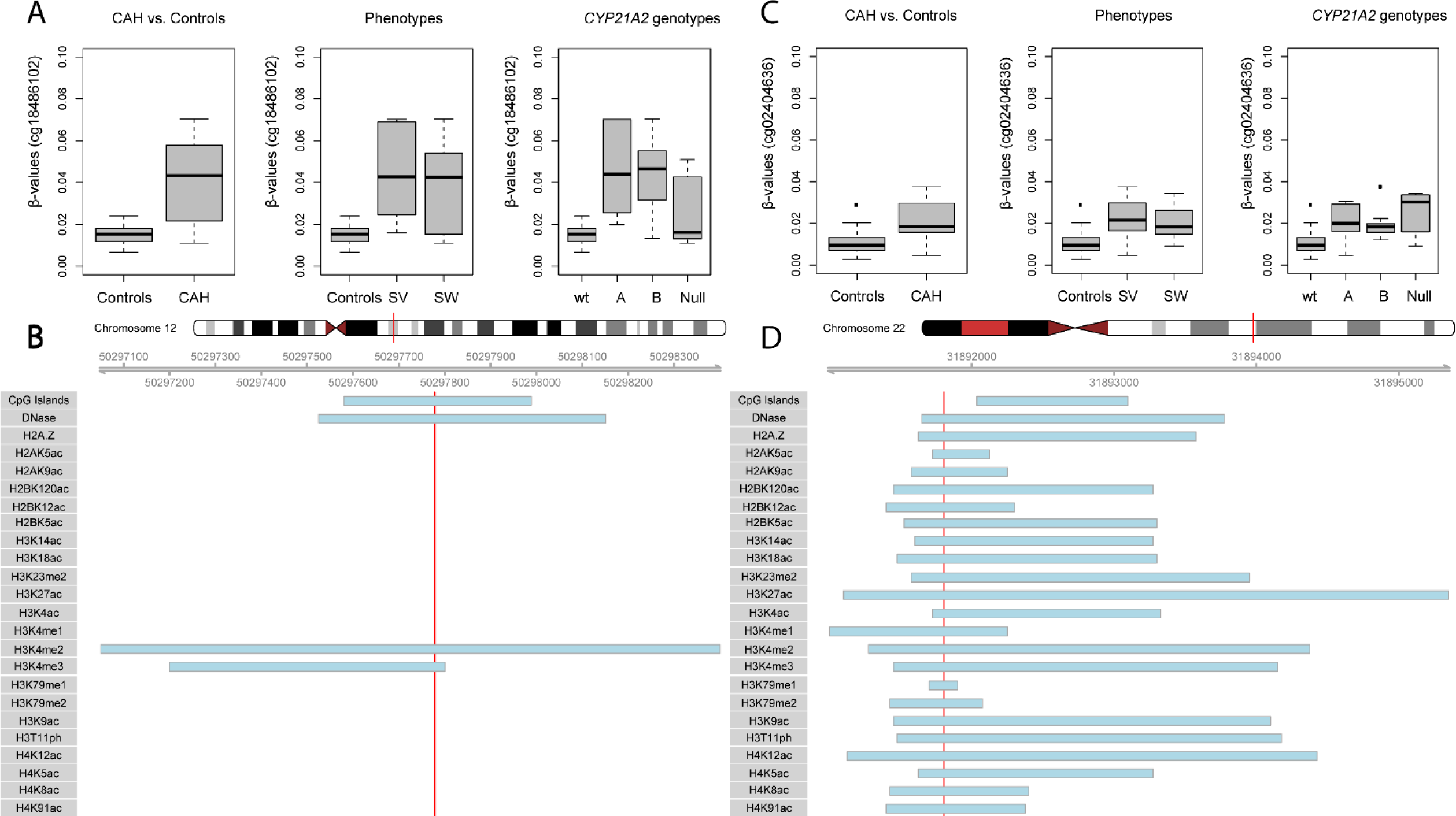
Level of methylation for differentially methylated probes and genomic locations with of significant CpG sites. Figures A and C presents β-values for cg18486102 (A) and cg02404636 (C) identified in the genome-wide analysis. Values are shown separately for all experimental groups. Figure B and D shows the epigenetic environment for cg18486102 (B) and cg02404636 (D). The red line indicates the location of the differentially methylated probes while blue bars denote enriched regions with specific epigenetic features. Three extreme outliers for cg18486102 and four for cg02404636 were removed in the visualisation (boxplots A and C).

The site cg18486102 is located in a CpG island in the transcriptional start site (TSS200) of the *FAIM2* gene on chromosome 12. It also overlaps with a DNase1 hypersensitive site and the epigenomic marks H3K4me2 and H3K4me3 (Figure 1B and D). Moreover, when relating β-values for cg18486102 to the daily dose of glucocorticoids, there was a significant interaction between dose (HC equivalent dose [mg/m2]) and sex (β=0.04, F(3,58)=2.1, *p*=0.02). The CpG site cg02404636 is located in the TSS1500 of the *SFI1* gene on chromosome 22. It also appeared to be located in a small epigenomic hotspot enriched for various histone modifications and a DNase1 hypersensitive site (Figure 1D). There was no association between dose and methylation for cg02404636. Genomic locations for the markers were acquired from the Roadmap Epigenomics Project using data from CD4+ T-cells (http://egg2.wustl.edu/roadmap/web_portal/).

### 3.3 Clinical outcome and association with DNA methylation

We identified higher levels of serum C-peptide in patients with CAH (β=0.2, F(4,56)=3.1, *p*=0.044). Additionally, levels of C-peptide and HbA1c were positively correlated with patient phenotype (*rho*=0.261, *p*=0.044 and *rho*=0.274, *p*=0.034) and genotype (*rho*=0.265, *p*=0.044 and *rho*=0.281, *p*=0.033), respectively (Table 1).

**Table 1.** Group comparison of CAH vs. controls: Correlations of genotype and phenotype status with cognitive and metabolic outcome. Patients with CAH had significantly higher C-peptide levels. Higher levels of C-peptide and HbA1c are associated with a more severe CAH genotype and phenotype. Significant findings are marked in boldface.

| **Growth**<br>**(Mean ±1SD)** | **CAH**<br>**(n=28)** | **C (n=34)** | **P [CAH]**<br>**CAH vs. C** | **P [CAH x**<br>**sex]**<br>**CAH vs. C** | **Rho**<br>**[genotype]** | **P**<br>**[genotype]** | **Rho**<br>**[phenotype]** | **P**<br>**[phenotype]** |
| --- | --- | --- | --- | --- | --- | --- | --- | --- |
| Height (cm) | 162.1 (12.8) | 160.2 (20.1) | 0.592 | 0.142 | -0.137 | 0.291 | -0.125 | 0.337 |
| Weight (kg) | 61.9 (16.4) | 56.3 (22.1) | 0.447 | 0.407 | -0.009 | 0.946 | 0.013 | 0.919 |
| BMI | 23.4 (5.0) | 20.9 (4.4) | 0.146 | 0.702 | 0.135 | 0.300 | 0.142 | 0.275 |
| **Glucose homeostasis (Mean ±1SD)** |  |  |  |  |  |  |  |  |
| B-Glucose (mmol/L) | 4.7 (0.3) | 4.7 (0.4) | 0.992 | 0.799 | -0.015 | 0.911 | -0.009 | 0.944 |
| S-Insulin (mIE/L) | 11.5 (5.3) | 8.8 (4.1) | 0.103 | 0.809 | 0.166 | 0.209 | 0.169 | 0.201 |
| S-C-peptide (nmol/L) | 0.8 (0.3) | 0.6 (0.2) | **0.044** | 0.591 | 0.261 | **0.044** | 0.274 | **0.034** |
| B-HbA<sub>1c</sub> (mmol/mol) | 31.9 (2.5) | 30.7 (2.2) | 0.052 | 0.700 | 0.265 | **0.044** | 0.281 | **0.033** |
| **Lipid profile (Mean ±1SD)** |  |  |  |  |  |  |  |  |
| P-Triglycerides (mmol/L) | 0.7 (0.3) | 0.7 (0.4) | 0.783 | 0.962 | 0.017 | 0.898 | 0.004 | 0.974 |
| P-Cholesterol (mmol/L) | 4.3 (0.9) | 4.2 (0.7) | 0.758 | 0.784 | 0.032 | 0.806 | 0.037 | 0.779 |
| P-HDL cholesterol (mmol/L) | 1.5 (0.5) | 1.5 (0.3) | 0.996 | 0.551 | -0.046 | 0.724 | -0.024 | 0.857 |
| P-LDL cholesterol (mmol/L) | 2.4 (0.8) | 2.4 (0.6) | 0.818 | 0.949 | 0.056 | 0.668 | 0.05 | 0.702 |
| LDL/HDL ratio | 1.7 (0.6) | 1.7 (0.7) | 0.740 | 0.878 | 0.052 | 0.69 | 0.031 | 0.812 |
| **Cognitive performance (Mean ±1SD)** |  |  |  |  |  |  |  |  |
| Coding (Sc) | 10.3 (2.6) | 10.5 (2.8) | 0.974 | 0.988 | -0.045 | 0.735 | -0.03 | 0.821 |
| Matrices (Sc) | 10.3 (3.6) | 10.6 (3.4) | 0.241 | 0.222 | -0.107 | 0.42 | -0.082 | 0.535 |
| Vocabulary (Sc) | 10.6 (2.5) | 10.2 (2.2) | 0.767 | 0.969 | -0.014 | 0.917 | -0.001 | 0.993 |
| Digit Span (Sc) | 9.0 (2.6) | 9.5 (2.3) | 0.960 | 0.447 | -0.108 | 0.414 | -0.115 | 0.386 |
| Stroop Int (T) | 53.5 (8.3) | 54.1 (6.5) | 0.389 | 0.347 | -0.057 | 0.67 | -0.051 | 0.704 |

Abbreviations: CAH, congenital adrenal hyperplasia; C, controls; BMI, body mass index; B, whole blood concentration; S, blood serum
concentration; P, blood plasma concentration; HDL, high density lipoprotein; LDL, low density lipoprotein; Sc, scaled scores. T, T scores. Bold text indicates $p$ -value $<0.05$ .

Using the β-values as predictors, we investigated the association between DNA methylation of cg18486102 (*FAIM2* gene) and cg02404636 (*SFI1* gene) and patient outcomes. Results from this analysis are presented in table 2. In general, no significant associations were identified, except for the methylation of CpG cg02404636, which showed a significant interaction with the sex of the participant and fasting plasma HDL cholesterol levels (β=2.4, F(4,57)=2.3, *p*=0.035).

**Table 2.** Associations between DNA methylation (β-values) and metabolic and cognitive outcome for the CpG sites (cg18486102 and cg02404636). CpG cg02404636 showed a significant interaction with sex of participant and higher plasma HDL cholesterol levels in girls. Significant findings are marked in boldface.

| **Growth**<br>**(Mean ±1SD)** | **cg18486102**<br>**(*p*)** | **cg18486102 x**<br>**sex (*p*)** | **cg02404636**<br>**(*p*)** | **cg02404636**<br>**x sex (*p*)** |
| --- | --- | --- | --- | --- |
| Height (cm) | 0.797 | 0.600 | 0.755 | 0.385 |
| Weight (kg) | 0.098 | 0.604 | 0.837 | 0.235 |
| BMI | 0.100 | 0.317 | 0.431 | 0.277 |
| **Glucose homeostasis**<br>**(Mean ±1SD)** |  |  |  |  |
| Glucose (mmol/L) (B) | 0.452 | 0.554 | 0.994 | 0.465 |
| Insulin (mIE/L) (S) | 0.473 | 0.925 | 0.202 | 0.178 |
| C-peptide (nmol/L) (S) | 0.458 | 0.642 | 0.953 | 0.579 |
| HbA<sub>1c</sub> (mmol/mol) (B) | 0.684 | 0.756 | 0.282 | 0.336 |
| **Lipid profile**<br>**(Mean ±1SD)** |  |  |  |  |
| Triglycerides (mmol/L) (P) | 0.359 | 0.739 | 0.798 | 0.436 |
| Cholesterol (mmol/L) (P) | 0.421 | 0.179 | 0.453 | 0.187 |
| HDL cholesterol (mmol/L) (P) | 0.301 | 0.499 | 0.661 | **0.035** |
| LDL cholesterol (mmol/L) (P) | 0.197 | 0.075 | 0.591 | 0.693 |
| LDL/HDL Ratio | 0.122 | 0.106 | 0.759 | 0.455 |
| **Cognitive performance**<br>**(Mean ±1SD)** |  |  |  |  |
| Coding (Sc) | 0.858 | 0.099 | 0.356 | 0.530 |
| Matrices (Sc) | 0.895 | 0.373 | 0.983 | 0.176 |
| Vocabulary (Sc) | 0.653 | 0.094 | 0.108 | 0.069 |
| Digit Span (Sc) | 0.910 | 0.127 | 0.245 | 0.342 |
| Stroop Int (Sc) | 0.113 | 0.066 | 0.572 | 0.978 |

Abbreviations: BMI, body mass index; B, whole blood concentration; S, blood serum
concentration; P, blood plasma concentration; HDL, high density lipoprotein; LDL, low density lipoprotein; Sc; scaled scores. Bold text indicates *p*-value <0.05.

## 4. Discussion

We investigated genome-wide DNA methylation in peripheral CD4+ T cells in patients with CAH to elucidate whether epigenomic reprogramming occurs. We also studied the association between patient outcome and DNA methylation. In addition, we investigated DNA methylation in a small group of patients with CAH who had been treated with DEX prenatally to identify if there is an imprinting effect of such therapy that may affect the long-term health. We choose the T cell because it is easily accessible and to minimise confounding effects from having a mix of cells with different methylation profiles. The T cell can be used to study the effects on this cell type *per se*, but also as a model system to study mechanisms or events that may occur in other cell types during embryogenesis and during postnatal development [40]. Moreover, we have previously observed broad associations between differences in DNA methylation in peripheral CD4+ T-cells between controls and individuals treated with DEX during the first trimester but not having CAH [27].

No significant associations were identified for either CAH or prenatal DEX treatment for T cell methylation in patients with CAH. This is contradictory in light of our previous study where we observed broad associations with DEX in the CD4+ T-cell methylome in CAH *unaffected* individuals [27]. In our previous study we identified broad differences between treated individuals and controls that were mainly associated with immune functioning and inflammation [27]. One reason for the lack of association with DEX in the present investigation could be the small sample size of CAH-DEX patients. On comparing methylation between CAH patients and controls, we did not observe a main effect of CAH *per se*. This finding is in itself important as this is a patient group with a lifelong chronic disorder that requires lifelong treatment and includes risks for several co-morbidities.

However, we did identify significant positive correlations with CAH phenotype and genotype in two CpG sites located in the TSS of two genes, *FAIM2* (cg18486102) and *SFI1* (cg02404636) (Figure 1A and C), where hypermethylation was associated with a more severe genotype and phenotype. Both CpG sites were located in regions overlapping with several epigenetic markers (Figure 1B and D). The CpG cg18486102 overlapped with a CpG island and a DNase1 hypersensitive site and with a region enriched for two histone modifications (H3K4me2 and H3K4me3). These two histone modifications are markers of actively transcribed promotors and transcription factor binding sites [41, 42]. The CpG cg02404636 also overlapped with a DNase1 hypersensitive site and was located near a CpG island. The differentially methylated CpG also overlapped with regions enriched with a large number of histone modifications (Figure 1D). Furthermore, investigating both cg18486102 and cg02404636 using Ensambl (http://grch37.ensembl.org/index.html) we observed that they were located in promoters active in CD4+ T cells.

Importantly, while cg18486102 was filtered based on FDR, it was among the top most significant CpG probes correlating with genotype (*rho*=0.54, p<0.0001, adjusted *p*=0.153) and the most significant probe in the comparison between patients and controls (*t*=4.86, *p*<0.0001, adjusted *p*=0.315). Furthermore, the finding that both CpGs seemed to be consistently significant, or among the top probes, for both phenotype and genotype is in line with the observation that there is a good genotype-phenotype correlation for CAH [2].

In the present cohort of patients with CAH we did not observe any cognitive deficits when compared with controls. However, patients with CAH showed significantly higher plasma concentrations of C-peptide. We also found positive correlations between C-peptide and HbA1c levels with the severity of patient genotype or phenotype (Table 1). These results indicate that patients with a more severe *CYP21A2* mutation and clinical phenotype may be more susceptible to insulin resistance. Thus, we provide evidence that the severity of the disorder is associated with affected glucose homeostasis consistent with the published evidence [7, 8, 12].

*FAIM2* is a widely expressed membrane-associated protein with very high expression in the brain compared to other tissues [43, 44]. It has been described as an anti-apoptotic protein protecting neurons from Fas ligand activated apoptosis [45, 46]. We suggest that hypermethylation of cg18486102 in the TSS of *FAIM2* could result in neurons being more sensitive to apoptosis, reductions in axonal growth, or both. In a case-control study of brain morphology in patients with CAH the authors identified widespread reductions in white matter structural integrity and reductions of volumes in several brain regions [47]. Moreover, SNPs in *FAIM2* have been associated with obesity and in another study of *FAIM2* promotor methylation, several CpG sites were significantly associated with dyslipidaemia [48-50].

These results are consistent with our hypothesis on metabolic abnormalities in patients with CAH. One plausible reason for not finding a significant association with any metabolic or cognitive measure could be the small sample size given that there are several *p*-values in the 0.05-0.1 range (Table 2). Our preliminary data indicate that there indeed is a relationship between brain structure and methylation in patients with CAH since we previously have identified a positive association between *FAIM2* methylation and surface area of the medial occipito-temporal and lingual sulcus [15]. This might be an indication that differential methylation in *FAIM2* could affect the developmental trajectory of the brain in patients with CAH. Moreover, we also identified a significant interaction between GC dose (HC equivalent dose [mg/m2]) and sex with higher doses predicting higher methylation.

SFI1 is a centrin-binding protein involved in centrosome maturation and cell cycle progression and mitosis and is ubiquitously expressed throughout the body [51-53]. The hypermethylation of the TSS region of *SFI1* could possibly (if the methylation leads to decreased expression) interfere with tissue development by interfering with cell differentiation and proliferation. We identified a significant interaction between CpG methylation of *SFI1* and sex of participant, with higher plasma concentrations of HDL cholesterol in girls. The mechanism as to how and why this gene would influence blood lipid levels, however, is not known and therefore the results should be interpreted with caution.

### 4.1 Limitations

Our study has some limitations. First, because our sample size is relatively small, it is possible that we failed to detect small effects, particularly for rate outcomes (false negative). This issue is in part supported by the finding that several *p*-values in the between group comparisons on metabolic and cognitive outcome and the association with methylation are between 0.05-0.1, suggesting that the lack of power could be attributed to small sample size. A potential confounding factor is that the study includes children, who may not develop any metabolic or cognitive sequelae until later in life, or it may simply be that there are no real differences in these data. However, our study benefits from a very unique cohort of patients from which data on a large array of variables have been acquired, including the genetic status of patients together with outcome parameters. This unfortunately also means that there is a lack of additional cohorts for validation at this point in time and limits the possibility for meaningful meta-analyses. Furthermore, we cannot infer any direct conclusions of hypermethylation in *FAIM2* and *SFI1* TSSs on gene expression because of the lack of RNA expression data, but we do know that TSS hypermethylation is typically associated with transcriptional silencing. In addition, the results are strengthened by the fact that both CpGs are located in promoters that are active in CD4+ T cells.

## 5. Conclusion

This is the first analysis of genome-wide DNA methylation in patients with CAH. We identified two CpGs (cg18486102 and cg02404636) located in the TSS region of the genes *FAIM2* (cg18486102) and *SFI1* (cg02404636) that were associated with the severity of the clinical CAH phenotype and *CYP21A2* genotype. In light of the long-term metabolic and cognitive sequelae patients with CAH are at risk of, both genes should be considered biologically relevant. Although we did not observe a direct association between methylation and metabolic or cognitive outcome, differential methylation in their respective TSS region may still have important implications for the long-term outcome in patients with CAH. Further studies on larger cohorts in addition to functional studies are needed to determine whether the identified alterations affect the health of the individual.

## Data Availability

all data are available upon request

## Acknowledgements

Not applicable.

## Abbreviations

CAH: congenital adrenal hyperplasia
SW CAH: salt-wasting
SV CAH: simple virilizing
DEX: dexamethasone
NC: non-classic
GC: glucocorticoid
B: blood
P: plasma
S: serum
HDL: high density lipoprotein
LDL: low density lipoprotein
WISC: Wechsler Intelligence Scale for Children
WAIS: Wechsler Adult Intelligence Scale
PBMC: peripheral blood mononuclear cells
MACS: magnetic activated cell sorting
450K array: Illumina Infinium Human Methylation450 BeadChip Array
SNP: single nucleotide polymorphism
FDR: false discovery rate
TSS: transcriptional start site.

## Notes

**Funding:** This work was supported by Marianne and Marcus Wallenberg Foundation, International Fund Congenital Adrenal Hyperplasia (IFCAH)/European Society for Pediatric Endocrinology (ESPE), Stockholm County Council (ALF-SLL), Stiftelsen Frimurare Barnhuset i Stockholm (SFBS), Svenska Läkaresällskapet, Stiftelsen Samariten, Jerringfonden, Sällskapet Barnavård and Wera Ekströms Stiftelse för pediatrisk forskning.

### Competing Interest Statement

The authors have declared no competing interest.

### Funding Statement

This work was supported by Marianne and Marcus Wallenberg Foundation, International Fund Congenital Adrenal Hyperplasia (IFCAH)/European Society for Pediatric Endocrinology (ESPE), Stockholm County Council (ALF-SLL), Stiftelsen Frimurare Barnhuset i Stockholm (SFBS), Svenska Läkaresällskapet, Stiftelsen Samariten, Jerringfonden, Sällskapet Barnavård and Wera Ekströms Stiftelse för pediatrisk forskning.

### Author Declarations

All relevant ethical guidelines have been followed and any necessary IRB and/or ethics committee approvals have been obtained.

Any clinical trials involved have been registered with an ICMJE-approved registry such as ClinicalTrials.gov and the trial ID is included in the manuscript.

## References

[1] P.W. Speiser, R. Azziz, L.S. Baskin, L. Ghizzoni, T.W. Hensle, D.P. Merke, H.F. Meyer-Bahlburg, W.L. Miller, V.M. Montori, S.E. Oberfield, M. Ritzen, P.C. White, S. Endocrine, Congenital adrenal hyperplasia due to steroid 21-hydroxylase deficiency: an Endocrine Society clinical practice guideline, J Clin Endocrinol Metab, 95 (2010) 4133–4160.

[2] M.I. New, M. Abraham, B. Gonzalez, M. Dumic, M. Razzaghy-Azar, D. Chitayat, L. Sun, M. Zaidi, R.C. Wilson, T. Yuen, Genotype-phenotype correlation in 1,507 families with congenital adrenal hyperplasia owing to 21-hydroxylase deficiency, Proc Natl Acad Sci U S A, 110 (2013) 2611–2616.

[3] M.G. Forest, H. Betuel, M. David, Prenatal Treatment of Congenital Adrenal Hyperplasia due to 21-Hydroxylase Deficiency: Update 88 of the French Multicentric Study., Endocrinology Research, 15 (1989) 277–301.

[4] S. Nimkarn, K. Lin-Su, M.I. New, Steroid 21 hydroxylase deficiency congenital adrenal hyperplasia, Pediatric clinics of North America, 58 (2011) 1281-1300, xii.

[5] P.C. White, P.W. Speiser, Congenital adrenal hyperplasia due to 21-hydroxylase deficiency, Endocrine reviews, 21 (2000) 245–291.

[6] P.W. Speiser, P.C. White, Congenital adrenal hyperplasia, The New England journal of medicine, 349 (2003) 776–788.

[7] W. Arlt, D.S. Willis, S.H. Wild, N. Krone, E.J. Doherty, S. Hahner, T.S. Han, P.V. Carroll, G.S. Conway, D.A. Rees, R.H. Stimson, B.R. Walker, J.M. Connell, R.J. Ross, E. United Kingdom Congenital Adrenal Hyperplasia Adult Study, Health status of adults with congenital adrenal hyperplasia: a cohort study of 203 patients, J Clin Endocrinol Metab, 95 (2010) 5110–5121.

[8] H. Falhammar, L. Frisen, A.L. Hirschberg, C. Norrby, C. Almqvist, A. Nordenskjold, A. Nordenstrom, Increased Cardiovascular and Metabolic Morbidity in Patients With 21-Hydroxylase Deficiency: A Swedish Population-Based National Cohort Study, J Clin Endocrinol Metab, 100 (2015) 3520–3528.

[9] L. Karlsson, A. Gezelius, A. Nordenstrom, T. Hirvikoski, S. Lajic, Cognitive impairment in adolescents and adults with congenital adrenal hyperplasia, Clin Endocrinol (Oxf), 87 (2017) 651–659.

[10] W.V. Browne, P.C. Hindmarsh, V. Pasterski, I.A. Hughes, C.L. Acerini, D. Spencer, S. Neufeld, M. Hines, Working memory performance is reduced in children with congenital adrenal hyperplasia, Horm Behav, 67 (2015) 83–88.

[11] J. Helleday, I.A. Bartfai, E.M. Ritzén, a.F. M., Generel Intelligence and Cognitive Profile In Women With Congenital Adrenal Hyperplasia (CAH), Psychoneuroendocrinoiogy, 19 (1994) 343–356.

[12] P. Sartorato, E. Zulian, S. Benedini, B. Mariniello, F. Schiavi, F. Bilora, G. Pozzan, N. Greggio, A. Pagnan, F. Mantero, C. Scaroni, Cardiovascular risk factors and ultrasound evaluation of intima-media thickness at common carotids, carotid bulbs, and femoral and abdominal aorta arteries in patients with classic congenital adrenal hyperplasia due to 21-hydroxylase deficiency, J Clin Endocrinol Metab, 92 (2007) 1015–1018.

[13] S.A. Berenbaum, K.K. Bryk, S.C. Duck, Normal intelligence in female and male patients with congenital adrenal hyperplasia, Int J Pediatr Endocrinol, 2010 (2010) 853103.

[14] M.L. Collaer, P.C. Hindmarsh, V. Pasterski, B.A. Fane, M. Hines, Reduced short term memory in congenital adrenal hyperplasia (CAH) and its relationship to spatial and quantitative performance, Psychoneuroendocrinology, 64 (2016) 164–173.

[15] A. Van’t Westeinde, L. Karlsson, M. Thomsen Sandberg, A. Nordenstrom, N. Padilla, S. Lajic, Altered Gray Matter Structure and White Matter Microstructure in Patients with Congenital Adrenal Hyperplasia: Relevance for Working Memory Performance, Cerebral cortex, (2019).

[16] V. Messina, L. Karlsson, T. Hirvikoski, A. Nordenstrom, S. Lajic, Cognitive function of children and adolescents with congenital adrenal hyperplasia: importance of early diagnosis, J Clin Endocrinol Metab, (2020).

[17] H. Engberg, A. Butwicka, A. Nordenstrom, A.L. Hirschberg, H. Falhammar, P. Lichtenstein, A. Nordenskjold, L. Frisen, M. Landen, Congenital adrenal hyperplasia and risk for psychiatric disorders in girls and women born between 1915 and 2010: A total population study, Psychoneuroendocrinology, 60 (2015) 195–205.

[18] H. Falhammar, A. Butwicka, M. Landen, P. Lichtenstein, A. Nordenskjold, A. Nordenstrom, L. Frisen, Increased psychiatric morbidity in men with congenital adrenal hyperplasia due to 21-hydroxylase deficiency, J Clin Endocrinol Metab, 99 (2014) E554–560.

[19] A. Nordenstrom, L. Frisen, H. Falhammar, H. Filipsson, G. Holmdahl, P.O. Janson, M. Thoren, K. Hagenfeldt, A. Nordenskjold, Sexual function and surgical outcome in women with congenital adrenal hyperplasia due to CYP21A2 deficiency: clinical perspective and the patients’ perception, J Clin Endocrinol Metab, 95 (2010) 3633-3640.

[20] L. Frisen, A. Nordenstrom, H. Falhammar, H. Filipsson, G. Holmdahl, P.O. Janson, M. Thoren, K. Hagenfeldt, A. Moller, A. Nordenskjold, Gender role behavior, sexuality, and psychosocial adaptation in women with congenital adrenal hyperplasia due to CYP21A2 deficiency, J Clin Endocrinol Metab, 94 (2009) 3432–3439.

[21] L. Wallensteen, M. Zimmermann, M. Thomsen Sandberg, A. Gezelius, A. Nordenstrom, T. Hirvikoski, S. Lajic, Sex-dimorphic effects of prenatal treatment with dexamethasone, J Clin Endocrinol Metab, (2016) jc20161543.

[22] P.D. Gluckman, M.A. Hanson, T. Buklijas, F.M. Low, A.S. Beedle, Epigenetic mechanisms that underpin metabolic and cardiovascular diseases, Nature Reviews Endocrinology, 5 (2009) 401.

[23] P.A. Jones, Functions of DNA methylation: islands, start sites, gene bodies and beyond, Nature reviews. Genetics, 13 (2012) 484-492.

[24] T. Ronn, P. Volkov, C. Davegardh, T. Dayeh, E. Hall, A.H. Olsson, E. Nilsson, A. Tornberg, M. Dekker Nitert, K.F. Eriksson, H.A. Jones, L. Groop, C. Ling, A six months exercise intervention influences the genome-wide DNA methylation pattern in human adipose tissue, PLoS genetics, 9 (2013) e1003572.

[25] M. Szyf, The early-life social environment and DNA methylation, Clinical genetics, 81 (2012) 341–349.

[26] S. Stringhini, S. Polidoro, C. Sacerdote, R.S. Kelly, K. van Veldhoven, C. Agnoli, S. Grioni, R. Tumino, M.C. Giurdanella, S. Panico, A. Mattiello, D. Palli, G. Masala, V. Gallo, R. Castagne, F. Paccaud, G. Campanella, M. Chadeau-Hyam, P. Vineis, Life-course socioeconomic status and DNA methylation of genes regulating inflammation, International journal of epidemiology, 44 (2015) 1320–1330.

[27] L. Karlsson, M. Barbaro, E. Ewing, D. Gomez-Cabrero, S. Lajic, Epigenetic Alterations Associated With Early Prenatal Dexamethasone Treatment, Journal of the Endocrine Society, 3 (2019) 250–263.

[28] D. Wechsler, WAIS-IV administration and scoring manual, San Antonio, TX: Psychological Corporation 2008.

[29] J. Donders, A short form of WISC-III for clinical use, Psychol Assess, 9 (1997).

[30] C.J. Golden, S.M. Freshwater, The Stroop color and word test-A manual for clinical and experimental uses, Chicago: Stoelting Co 1998.

[31] L.E. Reinius, N. Acevedo, M. Joerink, G. Pershagen, S.E. Dahlen, D. Greco, C. Soderhall, A. Scheynius, J. Kere, Differential DNA methylation in purified human blood cells: implications for cell lineage and studies on disease susceptibility, PLoS One, 7 (2012) e41361.

[32] L. Karlsson, M. Barbaro, E. Ewing, D. Gomez-Cabrero, S. Lajic, Epigenetic alterations associated with early prenatal dexamethasone treatment, Journal of the Endocrine Society, (2018) js.2018-00377-js.02018-00377.

[33] P. Du, W.A. Kibbe, S.M. Lin, lumi: a pipeline for processing Illumina microarray, Bioinformatics, 24 (2008) 1547–1548.

[34] P. Du, X. Zhang, C.C. Huang, N. Jafari, W.A. Kibbe, L. Hou, S.M. Lin, Comparison of Beta-value and M-value methods for quantifying methylation levels by microarray analysis, BMC Bioinformatics, 11 (2010) 587.

[35] J. Nordlund, C.L. Bäcklin, P. Wahlberg, S. Busche, E.C. Berglund, M. Eloranta, T. Flaegstad, E. Forestier, B. Frost, A. Harila-Saari, M. Heyman, Ó.G. Jónsson, R. Larsson, J. Palle, L. Rönnblom, K. Schmiegelow, D. Sinnett, S. Söderhäll, T. Pastinen, M.G. Gustafsson, G. Lönnerholm, A. Syvänen, Genome-wide signatures of differential DNA methylation in pediatric acute lymphoblastic leukemia, Genome Biology, 14 (2013) r105.

[36] F. Marabita, M. Almgren, M.E. Lindholm, S. Ruhrmann, F. Fagerstrom-Billai, M. Jagodic, C.J. Sundberg, T.J. Ekstrom, A.E. Teschendorff, J. Tegner, D. Gomez-Cabrero, An evaluation of analysis pipelines for DNA methylation profiling using the Illumina HumanMethylation450 BeadChip platform, Epigenetics, 8 (2013) 333–346.

[37] J.T. Leek, W.E. Johnson, H.S. Parker, A.E. Jaffe, J.D. Storey, The sva package for removing batch effects and other unwanted variation in high-throughput experiments., Bioinformatics, 28 (2012) 882–883.

[38] M.E. Ritchie, B. Phipson, D. Wu, Y. Hu, C.W. Law, W. Shi, G.K. Smyth, limma powers differential expression analyses for RNA-sequencing and microarray studies, Nucleic Acids Res, 43 (2015) e47.

[39] J. Li, R. Tibshirani, Finding consistent patterns: a nonparametric approach for identifying differential expression in RNA-Seq data, Statistical methods in medical research, 22 (2013) 519–536.

[40] B. Ma, E.H. Wilker, S.A. Willis-Owen, H.M. Byun, K.C. Wong, V. Motta, A.A. Baccarelli, J. Schwartz, W.O. Cookson, K. Khabbaz, M.A. Mittleman, M.F. Moffatt, L. Liang, Predicting DNA methylation level across human tissues, Nucleic Acids Res, 42 (2014) 3515–3528.

[41] B.E. Bernstein, M. Kamal, K. Lindblad-Toh, S. Bekiranov, D.K. Bailey, D.J. Huebert, S. McMahon, E.K. Karlsson, E.J. Kulbokas, 3rd, T.R. Gingeras, S.L. Schreiber, E.S. Lander, Genomic maps and comparative analysis of histone modifications in human and mouse, Cell, 120 (2005) 169–181.

[42] Y. Wang, X. Li, H. Hu, H3K4me2 reliably defines transcription factor binding regions in different cells, Genomics, 103 (2014) 222–228.

[43] A. Reich, C. Spering, K. Gertz, C. Harms, E. Gerhardt, G. Kronenberg, K.A. Nave, M. Schwab, S.C. Tauber, A. Drinkut, K. Harms, C.P. Beier, A. Voigt, S. Gobbels, M. Endres, J.B. Schulz, Fas/CD95 regulatory protein Faim2 is neuroprotective after transient brain ischemia, J Neurosci, 31 (2011) 225–233.

[44] S.C. Tauber, K. Harms, B. Falkenburger, J. Weis, B. Sellhaus, R. Nau, J.B. Schulz, A. Reich, Modulation of Hippocampal Neuroplasticity by Fas/CD95 Regulatory Protein 2 (Faim2) in the Course of Bacterial Meningitis, J Neuropathol Exp Neurol, 73 (2014) 2–13.

[45] C.P. Beier, J. Wischhusen, M. Gleichmann, E. Gerhardt, A. Pekanovic, A. Krueger, V. Taylor, U. Suter, P.H. Krammer, M. Endres, M. Weller, J.B. Schulz, FasL (CD95L/APO-1L) resistance of neurons mediated by phosphatidylinositol 3-kinase-Akt/protein kinase B-dependent expression of lifeguard/neuronal membrane protein 35, J Neurosci, 25 (2005) 6765–6774.

[46] M. Fernandez, M.F. Segura, C. Sole, A. Colino, J.X. Comella, V. Cena, Lifeguard/neuronal membrane protein 35 regulates Fas ligand-mediated apoptosis in neurons via microdomain recruitment, Journal of neurochemistry, 103 (2007) 190–203.

[47] E.A. Webb, L. Elliott, D. Carlin, M. Wilson, K. Hall, J. Netherton, J. Reed, T.G. Barrett, V. Salwani, J.D. Clayden, W. Arlt, N. Krone, A.C. Peet, A.G. Wood, Quantitative Brain MRI in Congenital Adrenal Hyperplasia: In Vivo Assessment of the Cognitive and Structural Impact of Steroid Hormones, J Clin Endocrinol Metab, 103 (2018) 1330–1341.

[48] G. Thorleifsson, G.B. Walters, D.F. Gudbjartsson, V. Steinthorsdottir, P. Sulem, A. Helgadottir, U. Styrkarsdottir, S. Gretarsdottir, S. Thorlacius, I. Jonsdottir, T. Jonsdottir, E.J. Olafsdottir, G.H. Olafsdottir, T. Jonsson, F. Jonsson, K. Borch-Johnsen, T. Hansen, G. Andersen, T. Jorgensen, T. Lauritzen, K.K. Aben, A.L. Verbeek, N. Roeleveld, E. Kampman, L.R. Yanek, L.C. Becker, L. Tryggvadottir, T. Rafnar, D.M. Becker, J. Gulcher, L.A. Kiemeney, O. Pedersen, A. Kong, U. Thorsteinsdottir, K. Stefansson, Genome-wide association yields new sequence variants at seven loci that associate with measures of obesity, Nat Genet, 41 (2009) 18–24.

[49] L. Paternoster, D.M. Evans, E.A. Nohr, C. Holst, V. Gaborieau, P. Brennan, A.P. Gjesing, N. Grarup, D.R. Witte, T. Jorgensen, A. Linneberg, T. Lauritzen, A. Sandbaek, T. Hansen, O. Pedersen, K.S. Elliott, J.P. Kemp, B. St Pourcain, G. McMahon, D. Zelenika, J. Hager, M. Lathrop, N.J. Timpson, G.D. Smith, T.I. Sorensen, Genome-wide population-based association study of extremely overweight young adults--the GOYA study, PLoS One, 6 (2011) e24303.

[50] L. Wu, X. Zhao, Y. Shen, M.X. Zhang, Y. Yan, D. Hou, L. Meng, J. Liu, H. Cheng, J. Mi, Promoter methylation of fas apoptotic inhibitory molecule 2 gene is associated with obesity and dyslipidaemia in Chinese children, Diabetes & vascular disease research, 12 (2015) 217–220.

[51] J.V. Kilmartin, Sfi1p has conserved centrin-binding sites and an essential function in budding yeast spindle pole body duplication, The Journal of cell biology, 162 (2003) 1211–1221.

[52] J. Martinez-Sanz, F. Kateb, L. Assairi, Y. Blouquit, G. Bodenhausen, D. Abergel, L. Mouawad, C.T. Craescu, Structure, dynamics and thermodynamics of the human centrin 2/hSfi1 complex, Journal of molecular biology, 395 (2010) 191–204.

[53] J. Martinez-Sanz, A. Yang, Y. Blouquit, P. Duchambon, L. Assairi, C.T. Craescu, Binding of human centrin 2 to the centrosomal protein hSfi1, The FEBS journal, 273 (2006) 4504–4515.

